# Using civil claim enquiry data to understand the context and impact of dog-related injuries in England and Wales between 2017 and 2024

**DOI:** 10.1101/2025.06.05.25329033

**Authors:** John Tulloch, Gemma Ahearne, Jasmine Moxey-Butler, James McNally

## Abstract

**Background:** Dog-related injuries, particularly bites, are a growing public health concern, yet key context for effective prevention remains limited. This study takes a novel approach by analysing civil claims enquiry data to describe the context and impact of dog bite and non-bite incidents in England and Wales.

**Methods:** Descriptive analysis of anonymised civil claims enquiry data from 2017 to 2024. Demographic and injury consequences were compared between bite and non-bite incidents using Chi2 and Mann-Witney U tests.

**Results:** A total of 816 incidents (842 claims) were analysed; 91.3% were dog bites, 6.7% dog strikes. Bites occurred outside of residential properties (40.9%) or on public highways or pavements (22.0%). Non-bite incidents were in public spaces (44.3%) or public highways or pavements (38.6%). Delivery workers accounted for 32.4% of victims. Most dogs were off lead at the time of injury (80.2% bites, 85.7% non-bites). Fractures occurred in 72.7% of non-bite incidents. Mental illness followed in 15.1% of bite cases and 10.0% of non-bites; 6.5% were diagnosed with a specific phobia, 4.1% with post-traumatic stress disorder. Work absence was reported in 59.5% of bites, whilst 54.3% reported lost earnings.

**Discussion:** Civil claims data offer valuable insights into the burden and context of dog-related injuries. Dog strikes, though less common, often result in severe injury. Most public incidents involve unrestrained dogs. This work provides emerging evidence of the psychological impact of dog-related injuries.

**Conclusions:** We present a novel methodology for contextual injury research and highlight the need to assess enforceable dog lead use on highways and certain public spaces.

**What is already known on this topic:** Dog-related injuries, particularly bites, are recognised as a growing public health concern. Contextual information critical for prevention strategies is limited, with a focus on injuries within the home. This study proposes a new methodology, of analysing legal claims data, to identify contextual injury information.

**What this study adds:** We now know that most public dog-related injuries occur when a dog is unrestrained. Dog strike-related injuries are likely to be more severe than previously thought. For the first time, we show the degree of psychological trauma resultant of these injuries.

**How might this study affect research, practice, or policy:** This study supports an exploration of legislative change to mandate lead usage in certain public spaces, reducing off-lead incidents and improving public safety. More research is needed into the psychological consequences of dog-related trauma and what patient support is needed.

## Background

The United Kingdom (UK) has a growing dog population; an estimated 12.6 million dogs in 2019 [1], and 13.6 million in 2024 [2]. Despite the benefits that dogs bring to society [3,4], there is always the potential risk of injury due to dog bites and strikes. Hospitalisations due to dog bites and strikes have risen across Great Britain [5–8]. In England they have increased from 4.76 hospital admissions per 100,000 population in 1998 to 18.7 in 2023 [5,6], whilst in Wales, they have risen from 16.3 per 100,000 in 2014 to 23.7 in 2022 [7]. Dog-related deaths in England and Wales averaged three per year between 2001 and 2021, showing little change over time [9].

Analysis of medical records provide quality demographic and injury related data. Young children have the highest incidence, yet the increase in national incidence is being driven by an increase in adult admissions [5,7,8,10]. These data have shown that injuries are more likely to occur in areas of higher socio-economic deprivation [5,8]. Injuries to adults are mainly to limb extremities, whilst to children they are to their head and necks [5,8]. Direct hospital health care costs have been estimated at a minimum of £70.8 million in England in 2017/18 and £2.2 million in Scotland in 2022 [5,8].

Medical records have contextual limitations. Since cases are identified through the ICD-10 Code ‘Bitten or stuck by dog’ it is challenging to differentiate between a dog bite and a dog strike [11]. Attempts have been made to differentiate, with estimates that 95% of child admissions and at least 77.5% of adult admissions are due to bites [5]. Incidence estimates likely underestimate the burden of dog-related injuries as people may attend emergency departments without being admitted, may attend a primary care physician or self-treat their injuries [12,13]. Hospital data have shown that more individuals are bitten at home than in any other setting [5,8]. It has been shown that many victims have been bitten while attempting to interact with a dog known to the victim [13,14]. Critically these data provide limited information about the events preceding an injury, nor the long-term health and life consequences of the injury. Effective injury prevention strategies require more contextual data. Unique data sources could offer deeper insights into how dog bites occur and their impact on victims.

In English and Welsh law The Animals Act 1971 makes the owner of a dog strictly liable for any injuries caused by the dog [15]. For liability of damage to be proven then the following statements must be proven; (1) the damage must be of a kind which unless the animal was restrained was likely to be severe, (2) the damage must have been caused by atypical characteristics for the species or those that appear only in certain situations, (3) the keeper must have known about these characteristics. To establish these criteria, solicitors gather extensive data on injury context, impact and subsequent ability to work. One could explore such cases by analysing those taken to civil court. They could be analysed, but would likely reflect more affluent defendants, as claims are only pursued when assets exist to cover damages and legal costs. Most solicitors’ firms will collect initial contextual and impact data before making a judgement on whether the case can be adopted. If these data were collated and analysed, they would remove the inherent socio-economic bias of analysing court records.

This study aimed to understand whether analysing civil liability claim enquiry data could provide substantial contextual information about dog bites and strikes to inform the development of effective intervention strategies.

## Methods

Anonymised civil claims enquiry data were provided by law firm Slee Blackwell Solicitors LLP who provide legal services throughout England and Wales [16]. They have a range of expertise and are seen as industry leaders in animal law [17]. This firm routinely collects personal injury enquiry data regarding dog-related incidents as part of their screening process for case adoption. The firm compiled such data from 1^st^ January 2017 to 31^st^ March 2024. They were subsequently cleaned, with sensitive and identifiable information removed. When fully anonymised by the firm they were securely shared with the research team.

These enquiry data contained information about injured person (IP) demographics (sex and age), incident details (date, location/land use [18], context of incident), dog details (breed, level of restraint), and consequences to IP (physical injuries, psychological injuries, medical treatment, absence from work and loss of earnings). Data were stratified between incidents involving dog bites or those that did not (for analytically purposes referred to as ‘non-bite incidents’).

Temporal and spatial trends of incidents were analysed descriptively. Sex and ages differences in bite or strike IPs were assessed using Chi^2^ and Mann-Witney U tests respectively. Incident and dog details were analysed descriptively. IP consequences were analysed descriptively, except a comparison between medical treatments between the two incident types, compared using Chi^2^ tests. Time off work and resultant loss of earnings were compared between incident types using Chi^2^ tests.

The data in this study were originally generated for legal purposes, not research. These data were provided to the legal firm following GDPR and the explicit consent that they could be shared to third parties. Data were cleaned and anonymised by the legal firm before sharing with the research team. This project therefore involves the secondary analysis of data and adheres to ethical principles of research integrity, data protection, and responsible use. As such the University of Liverpool Research Ethics team confirmed that no ethical approval was needed. All statistical analyses were carried out using R language (version 3.2.0; R Core Team 2015). The results were deemed statistically significant where P < 0.05.

## Results

A total 816 dog-related incidents (842 individual claims) were recorded. Most incidents involved one claimant (97.3%). Most incidents occurred in England (93.7%), with the South-East having the highest prevalence (19.2%) (Table S1). Incidents primarily involved dog bites (91.3%), 6.7% were dog strikes (Table 1).

**Table 1.** Injury mechanism in dog-related personal injury claim incidents (2017 – 2024)

| Injury mechanism | Percentage<br>(n=816) |
| --- | --- |
| **Involves a dog bite to a person** | **91.3%** |
| Bite | 89.7% |
| Bite and strike | 1.6% |
| **Not involving a dog bite to a person** | **8.7%** |
| Strike | 5.1% |
| Dog on dog attack<br>(no/minor human injury) | 1.7% |
| Pulled over/Fall/Trip | 1.0% |
| Other | 0.9% |

Seasonality of dog bites was identified, with 32.7% of incidents occurring during the summer (Fig. S1). No trend for day of the week was seen (Fig. S2). Incidents peaked within regular working hours (i.e. 9am-5pm) (Fig. S3).

Dog bite victims were mainly male (52.7%), whist non-bitten IPs were female (69.9%). Women were more than twice as likely to be involved in a non-bite incident than a bite incident compared to men (Table S2). There was a significant difference in age profile, with non-bite IPs (mean=50) being significantly older than bite IPs (mean=39) (Mann-Witney U=18568, p<0.001). Most individuals did not know the dog involved (80.3%) nor were aware of any previous incidents involving in the dog (89.5%).

Most dog bite incidents occurred on private residential properties (52.5%) (Table 2). The three most prevalent bite locations were; in front of a private residential property (22.4%), at the door of a private residential property (18.5%), and on a public highway or pavement (16.4%). Non-bite incidents mainly occurred in public spaces (44.3%). The most prevalent locales were; public highway or pavement (34.3%), outdoor recreational areas (28.6%), and ‘forestry, open land and water’ (8.6%).

**Table 2.**
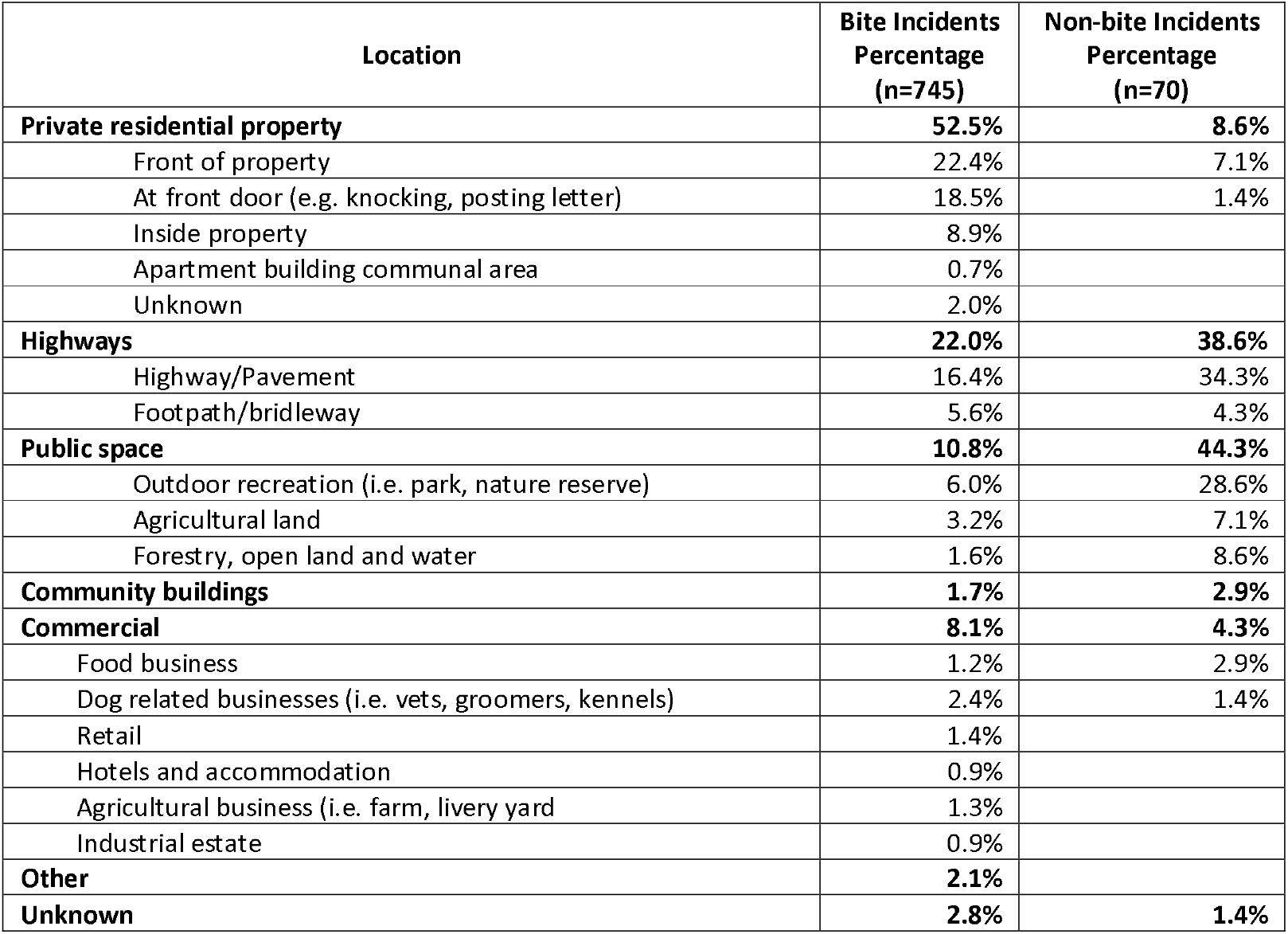
Locations of dog bite and non-bite incidents in dog-related personal injury claims.

Most IPs were injured when not at work; 52.4% of bite, and 90.0% of non-bite incidents (Table 3). Delivery workers made up almost a third of all bite incidents. The most common context for bites were: delivery workers entering the front of a property (22.4%); walking, exercising, playing in public without a dog (13.0%); and intervening in a dog-on-dog attack (10.7%). The common contexts for non-bite incidents were; walking, exercising, playing in public with own dog (44.3%); walking, exercising, playing in public without a dog (18.6%); and a dog escaping from a private property (18.6%).

**Table 3.**
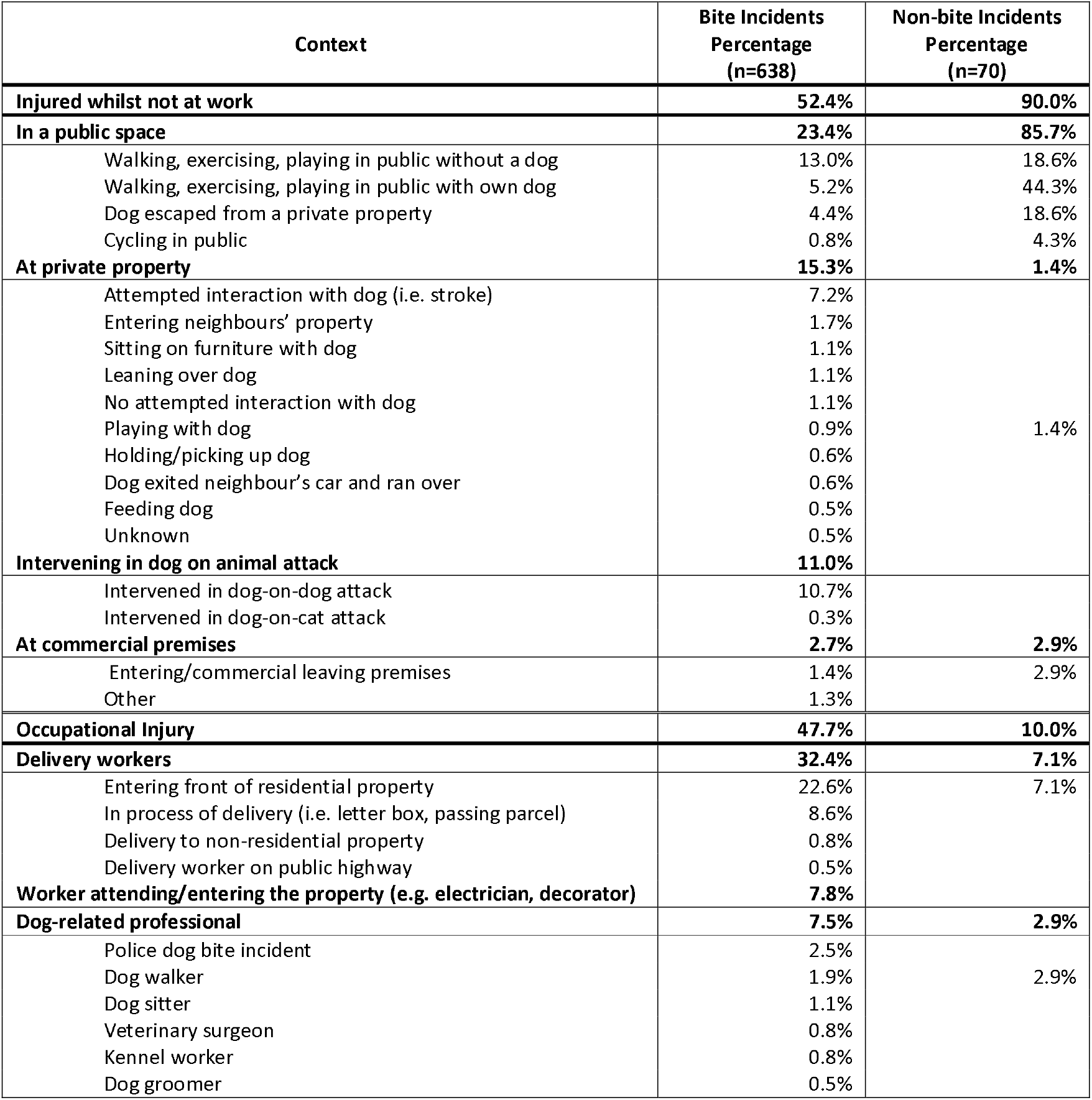
Context of dog bite and non-bite incidents in dog-related personal injury claims.

The majority of dogs involved in a bite (68.6%), or non-bite (77.5%) incident were reported to be with their owner. Most dogs were not restrained at the time of bite (80.2%) and non-bite incidents (85.7%). Incidents were reported to the police for 72.5% of bites, and 60.3% of non-bite incidents. Results about the dog’s breed, and their subsequent discussion, are found within the supplementary material.

Ninety-eight percent (97.9%) of bite and 78.1% of non-bite incidents resulted in a severe physical injury. For bites these were primarily ‘bite(s) wounds’ (46.7%), puncture wounds (39.7%), and lacerations (15.0%) (Table S4). Fractures amounted to 3.6% of injury, whilst tissue loss or amputations were 3.1%. For non-bite incidents these were primarily fractures (72.7%), muscle/tendon/ligament damage (9.1%), and soft tissue damage (9.1%). The predominant anatomy injured by bites were wrist and hands (33.0%), knee and lower legs (18.9%), and elbow and forearm (18.9%) (Table 4). One in seven bite injuries were to the head. Non-bite injuries occurred mainly to the knee and lower leg (47.4%), and shoulder and upper arm (26.3%). Over twelve percent of non-bite injuries resulted in tibial plateau fractures.

**Table 4.** Anatomy injured resultant of dog-related incidents recorded in personal injury claims.

|  | **Bite Claimants with a physical injury (n=737) \*** | **Non-bite Claimants with a physical injury (n=57) \*** |
| --- | --- | --- |
| Wrist and hand | 33.0% | 12.3% |
| Knee and lower leg | 19.9% | 47.4% # |
| Elbow and forearm | 18.9% | 10.5% |
| Hip and thigh | 17.0% | 7.0% |
| Head | 14.0% | 8.8% |
| Abdomen, lower back, lumber spine and pelvis | 7.5% | 10.5% |
| Shoulder and Upper Arm | 6.0% | 26.3% |
| Chest | 6.0% | 3.5% |
| Ankle and foot | 1.5% | 14.0% |
| Genitals | 0.9% |  |
| Neck | 0.7% |  |
| Unspecified | 1.4% |  |

\* Multiple anatomical area may have been injured during the incident

The majority of bite (90.3%) and non-bite (75.7%) IPs self-reported psychological injuries (Table 5). Fifteen percent of bitten IPs, and 10% of non-bitten IPs were provided with a clinical diagnosis of a mental illness as defined by the Diagnostic and Statistical Manual of Mental Disorders (DSM-5)[19]. Overall, 6.5% of IPs were diagnosed with a specific phobia and 4.1% were diagnosed with Post-Traumatic Stress Disorder (PTSD). The most prevalent symptoms for bitten and non-bitten IPs were phobia, anxiety, disturbed sleep, and avoidance.

**Table 5.** Psychological injury resultant of dog-related incidents recorded in personal injury claims.

|  | **Bite**<br>**Claimants**<br>**(n=735)** | **Non-bite**<br>**Claimants**<br>**(n=70)** |
| --- | --- | --- |
| **No psychological injury** | 9.7% | 24.3% |
| **Clinically given a psychiatric diagnosis** | 15.1% | 10.0% |
| Post-Traumatic Stress Disorder | 4.2% | 2.9% |
| Specific Phobia (Situational Type) | 3.7% | 2.9% |
| Specific Phobia (Animal Type) | 3.1% |  |
| Adjustment Disorder with Anxiety | 1.8% |  |
| Other Specified Trauma and Stressor Related Disorder | 1.0% |  |
| Major Depressive Disorder and Generalised Anxiety Disorder | 0.4% |  |
| Adjustment Disorder with Anxiety and Depressed Mood | 0.3% | 2.9% |
| Adjustment Disorder with Mixed Anxiety and Depressed Mood | 0.3% |  |
| Episodic Panic Disorder | 0.1% |  |
| Generalised Anxiety Disorder | 0.1% |  |
| Mixed anxiety and depressive disorder | 0.1% |  |
| Adjustment Disorder with Mixed Anxiety and Mood Disturbance |  | 1.4% |
| **Self-reported psychological symptoms without clinical diagnosis\*** |  |  |
| Phobic symptoms | 53.6% | 34.3% |
| Anxiety and stress related disorder | 27.3% | 15.7% |
| Insomnia/Disturbed sleep/Nightmares | 21.1% | 18.6% |
| Avoidant symptoms | 18.8% | 20.0% |
| Self-image issues | 4.4% | 2.9% |
| PTSD symptoms | 2.7% | 2.9% |
| Panic attacks | 1.9% | 1.4% |
| Depression | 1.8% | 4.3% |
| Intrusive thoughts | 1.2% |  |
| Adjustment disorder | 0.8% |  |
| Emotional instability | 0.7% | 1.4% |
| **Unspecified** | 3.3% | 1.4% |

\*Multiple symptoms may have been experienced by an injured person

The majority of dog bite (86.0%) and non-bite (74.0%) IPs reported that the physical injury sustained resulted in hospital attendance. A quarter of bitten IPs (25.9%) and 30.2% of non-bitten IPs required surgical treatment. Few IPs mentioned length of stay within the hospital. Thirty bitten IPs had a median length of stay of 3 (range:2-7), whilst eight non-bitten IPs had a median stay of 3.5 (range:1-11). Thirty-one percent (31.0%) of bitten IPs visited their primary care physician, whilst 26.3% of non-bitten IPs did. A minority of IPs required ongoing treatment (13.6% of bitten, 43.3% of non-bitten); non-bitten IPs were more than four times more likely to need ongoing treatment than bitten IPs (OR=4.83, 95%CI2.75-8.43, p<0.001) (Table S5).

Of the claimants still working when the injury took place, 59.5% of bite and 56.1% of non-bite incidents were absent from work (Table S6). Few reported the length of time off work; the maximum recorded for a bitten IP was 5 years, and for a non-bitten IP was 5 months. Over half of bitten IPs (54.3%) and 41.4% of non-bitten IPs reported a loss of earning resultant of their injuries. Significantly more bitten IPs reported a loss of earnings (OR=1.68, 95% CI 1.02-2.79, p=0.04).

## Discussion and Conclusions

We have shown that civil claim enquiries data are a viable data source to explore the context and consequences of dog-related injuries. This novel data source offers the potential for research for other causes of injury. These data strongly implicate dogs off-lead in public spaces as a major inciting factor for dog-related injuries, and injury prevention strategies need to explore how lead use can be effectively legislated.

Most claimants reported that the dogs involved were with their owners and off lead. Given that half of bite incidents and over 90% of non-bite incidents occurred in public spaces, this raises concerns about the level of control owners maintain over off-lead dogs in these environments. National legislation concerning lead control is limited. The Countryside and Rights of Way Act 2000 permits dogs on open access land if it is on a short fixed-length lead (<2 metres) between March and July and always around livestock [20]. This has no impact on public highways or urban green spaces, where most injuries are occurring. The Highway Code advices that dogs should be ‘*Kept on a short lead when walking on the pavement, road or path shared with cyclists or horse riders*’ [21]. This is solely guidance, not law. Local authorities can introduce Public Space Protection Orders (PSPOs), under the Anti-Social Behaviour, Crime and Policing Act 2014, Sections 59 [22], to manage dogs out of control and gain the ability to fine those violating the orders. Some authorities apply PSPOs to state that dogs should be on a lead in town centres [23], cemeteries or churchyards [24,25], car parks [24], sports grounds and fields [23–25], nature reserves [23,24], and any roads (pavements, footways and verges) [23–25]. Most PSPOs do not specify that a lead needs to be fixed-length, and lead length is inconsistently defined, some omit it entirely [23], while one allows up to 5 metres [25], a length that limits effective control. It is unknown how well PSPOs are enforced, or how effective a deterrent they are. Despite PSPO’s existence, dog-related injuries in public places persist, suggesting a need to evaluate their effectiveness. We make recommend that national legislation is updated so that all dogs should be on a fixed-length short lead (<2 metres) on public highways and in urban green spaces (unless a local authority provides provisions for off-lead areas, or make areas exempt). This exemption provision is to ensure that the important balance between public safety and dog welfare can be achieved. This should be partnered with a nationally coordinated public communication campaign. It should be trialed regionally first to test effectiveness and identify any barriers to implementation. If implemented, we estimate these measures could potentially prevent up to 28% of public dog bites and 67.2% of non-bite incidents.

Personal/recreational injuries mainly occurred when utilising a public space invariably when the biting dog was off a lead. Occupational injuries were almost exclusively to delivery workers, who were injured at the front of people’s properties, mirroring prior analysis of occupational dog bite injuries [26]. Dogs biting delivery workers is often trivialised in society, yet we can see that these injuries are frequently life changing with loss of earnings, on-going physical health problems, and significant mental health issues. With only 1 in 3 delivery workers in direct employment [27], it is important that casual workers are protected from the everyday hazard of dogs. The onus of this should be on the agency and delivery company, but responsibility should also be placed on the dog owner. A front-door should not be opened with a dog present, gates and fences should be dog-secure, and internal letter cages or external letter boxes installed.

Individuals bitten by dogs were bitten more frequently during working hours and during the summer months, matching previous research [5,26,28,29]. Similarly the demographics and resultant injuries of IPs are consistent with the literature [5,8,10,26,28]. The majority of bitten individuals required hospital care, and a quarter required surgical treatment. A few required ongoing treatment, yet most were absent from work to some degree. These combined with the mental health consequences to IPs create a significant economic burden both to the individual and society.

Dog strike injuries primarily occurred in public spaces, whilst exercising, and a concerning number were injured by escaped dogs. These injuries occur when a dog, not adequately controlled by its owner, collides with an individual, often unexpectedly. The common resultant trauma of lower leg fractures reflects the high-energy injury mechanism of a dog strike [30,31]. The demographics of those injured, predominately women over 50 years old, are at heightened risk of fractures due to menopausal changes that can lead to low bone mass or osteoporosis [32,33]. Any fracture is of concern for this population as it increases the likelihood of future fractures [32,33]. The high rate of tibial plateau fractures was concerning. These require major surgery, which will take around 5 months to heal, 7% of elderly patients require a second operation, and 23% develop post-traumatic osteoarthritis [34]. These findings are reflected in our work where almost half of IPs required ongoing treatment. These injuries could be prevented through the regulated use of short leads.

The physical impact of dog-related injuries is well documented, but not so the psychological impact. Existing research focuses on children, with common consequences being PTSD, phobia, nightmares, flashbacks, anxiety, and social withdrawal [35,36]. We are unaware of any prior study stating the prevalence of psychological consequences of dog bites in a predominately adult population. At least one in six IPs received a clinical psychiatric diagnosis most commonly specific phobias and PTSD. The global annual prevalence of specific phobia is 5.5%, with the lifetime prevalence of 3.8% for the animal subtype [37]. The prevalence within our study exceeds these and show that dog-related injuries are a major risk to developing a specific phobia. This condition severely impairs individuals role in society, with 18.7% of individuals having their home, work, social and relationships impacted, with a resulting lower quality of life [37,38]. The prevalence of PTSD in this study is higher than that of global lifetime prevalence (3.9%) [39], highlighting that dog bites do pose a significant risk of PTSD. Half of those diagnosed with PTSD experience persistent symptoms [39,40]. Many respondents reported symptoms consistent with mental health disorders despite lacking a formal diagnosis, suggesting true prevalence may be higher. If representative, these findings underscore the need for a multi-disciplinary response, including improved recognition and referral pathways in emergency and primary care. Public awareness must also grow to reduce stigma and promote support for those affected by dog-related injuries.

The data source of this analysis has limitations. The majority of IPs did not know the dog, nor were they injured at home, whilst most research indicates that dogs are known to the victim and occur at home [5,8,13,29]. This is unsurprising as most individuals would not litigate against family members. These data are based on a single firm, and we do not know how representative they are. It is likely that these data only represent the more serious injuries caused by dogs, as it is unlikely that a civil claim is sought when it is a minor injury.

Analysis of civil claims enquiry data offer a novel approach to exploring the context of injuries. It has highlighted that dog-related injuries cause not only significant physical trauma, but also mental trauma, and that dog-strike injuries can be highly traumatic. We have shown that delivery workers are an occupational group at risk that require collective societal protection to minimise their likelihood of injuries. Most importantly we have shown that dog-related injuries in public spaces are almost all caused by dogs off lead. We recommend an exploration in the enforcement of lead usage in certain public locations.

## Data Availability

All data produced in the present study are available upon reasonable request to the authors

## Data availability statement

Data are available upon reasonable request.

## Contribution

Conceptualisation: JT, JM. Funding acquisition: JT, GA, JM. Methodology: JT, JM. Formal Analysis: JT. Writing original draft: JT. Writing review & editing: All authors Funding: This work was funded by both the Biotechnology and Biological Sciences Research Council [grant number BB/X511225/1] and Economic and Social Research Council [grant number ES/X004910/1] Impact Acceleration Accounts Grants.

## Competing interests

JM and JMB are current employees of Slee Blackwell Solicitors LLP

## Patient and public involvement

Patients and/or the public were not involved in the design, or conduct, or reporting, or dissemination plans of this work.

## Acknowledgements

We would like to thank all the staff at Slee Blackwell limited who compiled and cleaned the data. We would also like to thank all injured persons whose enquiry data were used in this analysis.

## Notes

### Author Declarations

The data in this study were originally generated for legal purposes, not research. These data were provided to the legal firm following GDPR and the explicit consent that they could be shared to third parties. Data were cleaned and anonymised by the legal firm before sharing with the research team. This project therefore involves the secondary analysis of data and adheres to ethical principles of research integrity, data protection, and responsible use. As such the University of Liverpool Research Ethics team confirmed that no ethical approval was needed.

