## Supplementary Material for "Using civil claim enquiry data to understand the context and impact of dog-related injuries in England and Wales between 2017 and 2024"

**Table S1.** Dog-related personal injury claim incidents stratified by region.

| Region | Percentage (n=809) |
|---|---|
| **England** | **93.7%** |
| South-East | 19.2% |
| South-West | 14.7% |
| East | 11.4% |
| West Midlands | 10.1% |
| London | 9.9% |
| North-West | 9.9% |
| Yorkshire and Humber | 7.9% |
| East Midlands | 6.2% |
| North-East | 3.6% |
| **Wales** | **6.3%** |

### Temporal Analysis

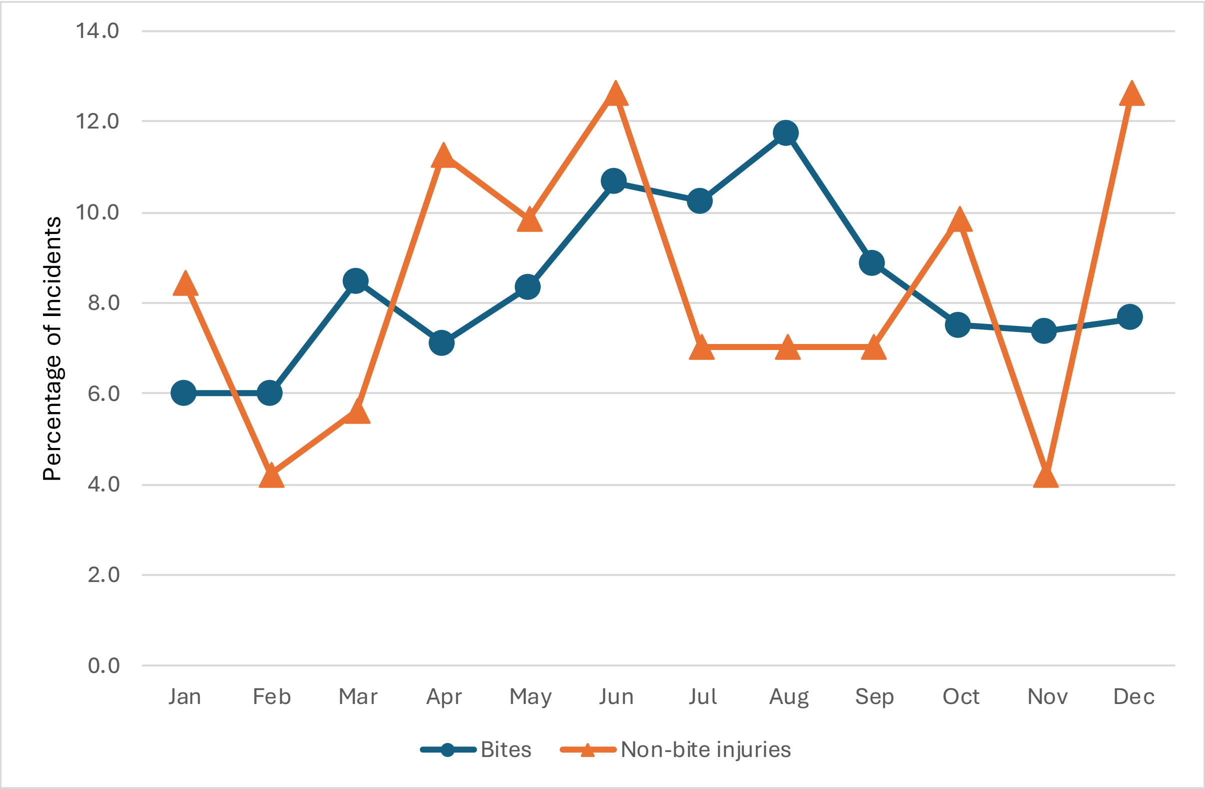


**Figure S1**. Seasonality of dog bite (n = 732) and non-bite incidents (n = 71) in dog-related personal injury claims incidents (2017-2023)
**Note:** Data for the year 2024 was not included as it did not include a full year of data.

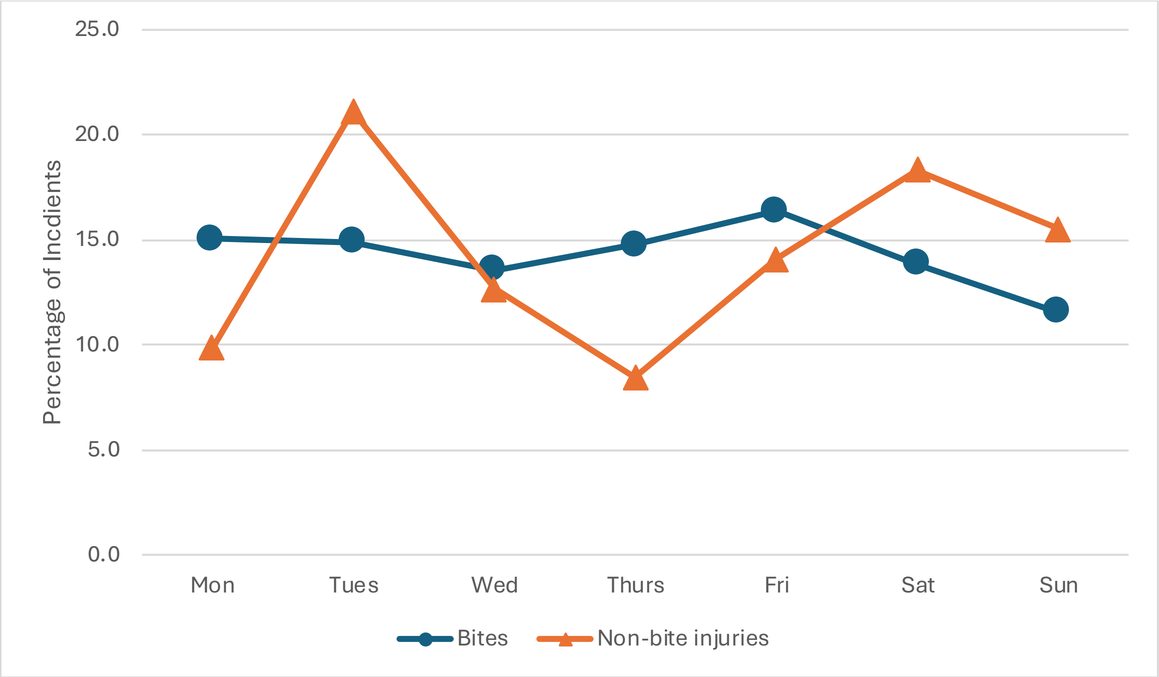


**Figure S2.** Day of the week analysis for dog bite (n = 745) and non-bite incidents (n = 71) in dog-related personal injury claims incidents

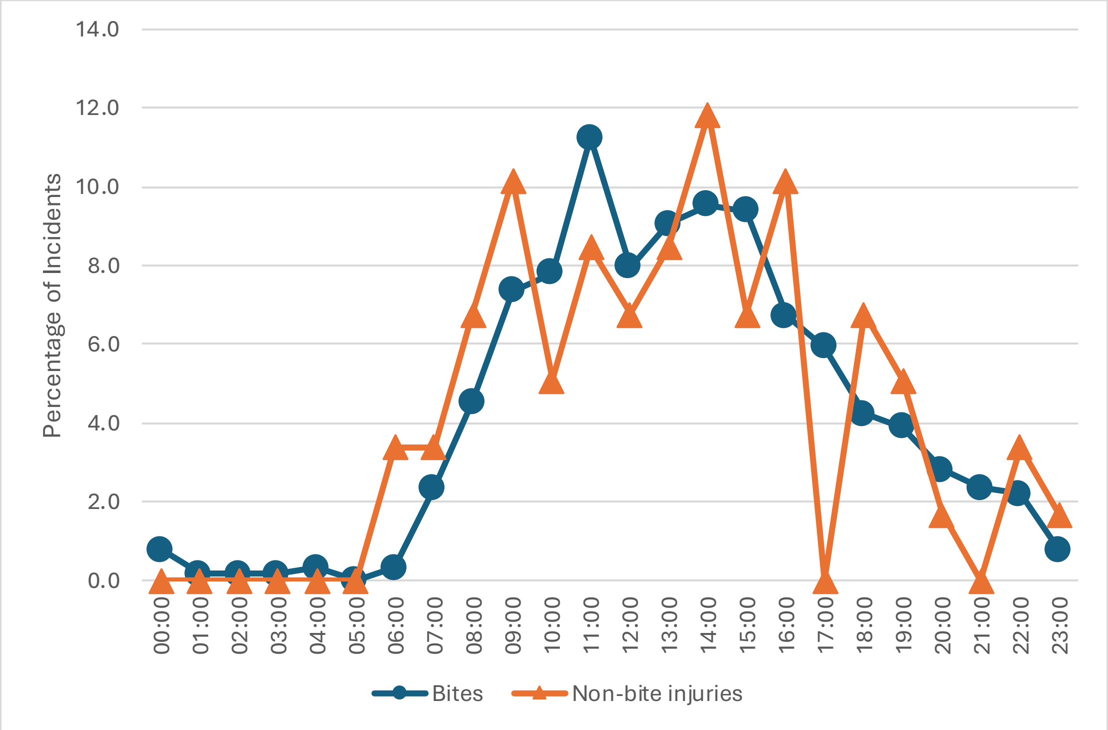


**Figure S3**. Time of day analysis for dog bite (n = 640) and non-bite incidents (n = 59) in dog-related personal injury claims incidents

### Injured Person Demographics

**Table S2.** Injured person demographics and knowledge of dog in dog-related personal injury claim incidents (2017 – 2024)

| | All incidents | Dog bite incidents | Non-bite incidents | Odds Ratio (95% CI) | p-value |
|---|---|---|---|---|---|
| **Sex** | n=842 | n=769 | n=73 | | |
| Female | 50.7% | 47.3% | 69.9% | | |
| Male | 49.3% | 52.7% | 30.1% | 0.39 (0.23-0.65) | <0.001 |
| **Age** | n=842 | n=769 | n=73 | | |
| Mean (range) | | 39 (1-89) | 50 (2-85) | N/A | |
| 1-20 | 12.8% | 13.3% | 8.2% | | |
| 21-30 | 16.5% | 17.4% | 6.8% | | |
| 31-40 | 21.5% | 22.5% | 11.0% | | |
| 41-50 | 18.5% | 18.1% | 23.3% | | |
| 51-60 | 18.9% | 18.5% | 23.3% | | |
| Over 60 | 11.8% | 10.3% | 27.4% | | |
| **Is the dog known to you?** | n=837 | n=764 | n=73 | | |
| Yes | 19.7% | 18.8% | 28.8% | | |
| No | 80.3% | 81.2% | 72.1% | 0.58 (0.34-1.00) | 0.05 |
| **Were they aware of previous incidents involving this dog?** | n=813 | n=746 | n=67 | | |
| Yes | 10.5% | 10.1% | 15.9% | 0.64 (0.32-1.37) | 0.23 |
| No | 89.5% | 88.9% | 84.1% | | |

**Dog breeds and management**

For 79.3% of the bite incidents, breed details of the dog could be provided. Of the 602 dogs involved, 69.4% were a recognised breed with the remainder being cross breeds (Table S3). The most prevalent dogs were; German Shepherds (15.6%), unknown mixed breeds (6.1%), Staffordshire Bull Terriers (5.5%), Border Collies (4.5%), and Rottweilers (4.5%). Only two dog types listed under the Dangerous Dogs Act 1991 were stated and were infrequently reported; American XL Bully (0.7%), American Pit Bull Terrier (0.5%). Regarding non-bite incidents, 56 incidents provided breed details for 61 dogs involved, 78.6% were a recognised breed. The most prevalent dogs were; Labradors (17.9%), German Shepherds (14.3%), Greyhounds (8.9%), Great Danes (5.4%), and Staffordshire Bull Terriers (5.4%).

The majority of breeds described by bitten IPs are broadly reflective of the prevalence of dog breeds within England and Wales (1,2), similar to previous UK research (3). Only one percent of dogs were described as a banned breed as defined by the Dangerous Dogs Act 1991 (4). This indicates that dog breeds do not have a strong correlation with bite incidence, and that bites are more likely to be associated with the degree and quality of training of the dog, the responsibleness of the owner, and any behavioural traits of the dog. However, non-bitten injuries (predominately strikes) are associated with larger dogs (i.e. Labradors, German shepherds, and greyhounds). These dogs often weigh more than 25kg, and impact at speed by these dogs is likely to lead to severe injury. These data must be treated with a slight degree of caution as it is known that the public find it hard to recognise breeds (5).

**Table S3.** Dog breeds involved in dog-related personal injury claim incidents (2017 – 2024)

| Breed (B) or Cross-breed (X) | Dog Breed | Bite Incidents Percentage (n=602) | Non-bite Incidents Percentage (n=56) |
|---|---|---|---|
| B | German Shepherd* | 15.6% | 14.3% |
| X | Unknown mixed breed | 6.1% | |
| B | Staffordshire Bull Terrier | 5.5% | 5.4% |
| X | Terrier cross | 5.0% | |
| B | Border Collie | 4.5% | |
| B | Rottweiler | 4.5% | 3.4% |
| B | Jack Russell Terrier | 3.7% | 3.4% |
| B | French Bulldog | 2.8% | |
| X | German Shepherd cross | 2.7% | 1.8% |
| B | Labrador | 2.7% | 17.9% |
| X | Spaniel cross | 1.7% | 1.8% |
| B | Alaskan Malamute | 1.5% | |
| B | Dachshund | 1.5% | |
| B | Husky | 1.5% | |
| X | Mastiff Cross | 1.5% | 1.8% |
| B | Bulldog | 1.3% | |
| X | Collie Cross | 1.3% | |
| B | Doberman | 1.3% | |
| B | Greyhound | 1.3% | 8.9% |
| X | Lurcher | 1.3% | |
| B | Akita | 1.2% | 3.4% |
| B | Belgian Malinois | 1.2% | |
| X | Bull dog cross | 1.2% | |
| B | Bull Mastiff | 1.2% | |
| B | Cocker Spaniel | 1.0% | |
| B | Springer Spaniel | 1.0% | |
| X | Staffordshire Bull Terrier Cross | 1.0% | 1.8% |
| X | Cockapoo | 0.8% | |
| X | St Bernard | 0.8% | |
| X | Akita cross | 0.7% | |
| B | American Bulldog | 0.7% | 3.4% |
| X | American XL Bully | 0.7% | 1.8% |
| B | Beagle | 0.7% | |
| B | Border Terrier | 0.7% | |
| B | Cane Corso | 0.7% | 1.8% |
| B | Dogue De Bordeaux | 0.7% | |
| X | Labradoodle | 0.7% | 1.8% |
| X | Labrador Cross | 0.7% | |
| B | American Pit Bull Terrier | 0.5% | |
| B | Boxer | 0.5% | |
| B | Chow Chow | 0.5% | |
| B | English Bulldog | 0.5% | |
| X | English Bull Terrier Cross | 0.5% | |
| B | Hungarian Vizsla | 0.5% | |
| X | Pocket Bully | 0.5% | |
| B | Weimaraner | 0.5% | |
| B | West Highland Terrier | 0.5% | |
| B | Yorkshire Terrier | 0.5% | |
| B | Airedale Terrier | 0.3% | |
| X | American Bull Dog Cross | 0.3% | 3.4% |
| B | Boston Terrier | 0.3% | |
| X | Doberman Type | 0.3% | |
| B | Golden Retriever | 0.3% | 1.8% |
| X | Golden Retriever Cross | 0.3% | |

| | | | |
|---|---|---|---|
| B | Great Dane | 0.3% | 5.4% |
| X | Great Dane Cross | 0.3% | 1.8% |
| X | Husky Cross | 0.3% | 3.4% |
| B | Irish Wolfhound | 0.3% | |
| B | Old English Sheepdog | 0.3% | |
| X | Pointer type | 0.3% | |
| B | Poodle | 0.3% | 3.4% |
| X | Retriever cross | 0.3% | |
| B | Rhodesian Ridgeback | 0.3% | |
| B | Scottish Terrier | 0.3% | |
| X | Shih Tzu Cross | 0.3% | |
| B | Tibetan Mastiff | 0.3% | |
| B | Whippet | 0.3% | |
| B | Anatolian Shepherd | 0.2% | |
| B | Basenji | 0.2% | |
| B | Belgian Shepherd | 0.2% | |
| B | Borzoi | 0.2% | |
| X | Boxer Cross | 0.2% | |
| B | Cairn Terrier | 0.2% | |
| B | Caucasian Shepherd | 0.2% | |
| B | Central Asian Shepherd | 0.2% | |
| B | Chihuahua | 0.2% | |
| B | Chinese crested | 0.2% | |
| B | Corgi | 0.2% | |
| X | Coonhound cross | 0.2% | |
| B | Dalmatian | 0.2% | 1.8% |
| B | Dutch Herder | 0.2% | |
| B | English Bull Terrier | 0.2% | |
| X | English pointer cross | 0.2% | |
| B | French Mastiff | 0.2% | |
| B | German Short Haired Pointer | 0.2% | 1.8% |
| B | Irish Terrier | 0.2% | |
| B | Kangal Shepherd | 0.2% | |
| X | Korean Jindo Cross | 0.2% | |
| B | Lakeland Terrier | 0.2% | |
| B | Large Munsterlander | 0.2% | |
| X | Leonburger cross | 0.2% | |
| B | Maremma Sheepdog | 0.2% | |
| X | Newfoundland cross | 0.2% | |
| B | Patterdale Terrier | 0.2% | |
| B | Polish Lowland Sheepdog | 0.2% | |
| B | Pomeranian | 0.2% | |
| X | Pomeranian cross | 0.2% | |
| X | Puggle | 0.2% | |
| X | Rottweiler cross | 0.2% | |
| B | Schnauzer (standard) | 0.2% | |
| B | Shar Pei | 0.2% | |
| B | Shih Tzu | 0.2% | |
| B | Swiss Shepherd | 0.2% | |
| X | Wolf crossbreed | 0.2% | |
| B | Australian Shepherd | 0.2% | 1.8% |
| X | Basset Hound Cross | 0.2% | 1.8% |

*This includes 12 police dogs

### Injured Person Consequences

**Table S4.** Injuries resultant of dog bite and non-bite incidents in dog-related personal injury claims

| Injury type | Bite Incidents Percentage* (n=748) | Non-bite Incidents Percentage* (n=55) |
|---|---|---|
| Bite(s) wound | 46.7% | |
| Puncture wound(s) | 39.7% | |
| Laceration(s) | 15.0% | |
| Fracture | 3.6% | 72.7% |
| Nerve damage | 3.5% | 3.6% |
| Muscle/tendon/ligament damage | 2.9% | 9.1% |
| Scratch(es) | 2.5% | 1.8% |
| Tissue loss (e.g. part of lip/tip of nose/ear) | 2.0% | |
| Abrasion | 1.2% | 3.6% |
| Cut(s) | 1.2% | 3.6% |
| Damaged nail/nail bed/nail loss | 1.2% | |
| Haematoma | 1.2% | 1.8% |
| Traumatic amputation of finger | 1.1% | |
| Soft tissue injury | 0.9% | 9.1% |
| Dental damage | 0.7% | 3.6% |
| Crush injury | 0.7% | |
| Sprain | 5.3% | 5.5% |
| Concussion | | 3.6% |
| Dislocation | | 3.6% |
| Other | 2.0% | |

* Multiple injuries may have occurred during the same incident

**Table S5** On-going treatments resultant of dog-related incidents recorded in personal injury claims

| Treatment | Bite Claimants (n=675)* | Non-bite Claimants (n=60)* |
|---|---|---|
| Physiotherapy | 4.6% | 31.7% |
| Counselling | 4.0% | 5.0% |
| Cosmetic surgery | 3.4% | |
| Cognitive Behavioural Therapy (CBT) | 2.4% | 1.7% |
| Eye movement desensitization and reprocessing (EMDR) | 0.9% | |
| Chiropractor | 0.3% | |
| Hospital outpatient services | 0.3% | 3.3% |
| Dentist | 0.1% | |
| Acupuncture | 0.1% | |
| No | 86.4% | 56.6% |

*Multiple symptoms may have been experienced by an injured person

**Table S6** – Impact of dog-related injuries on work absence and loss of earnings

| | Bite Claimants (n=676) | Non-bite Claimants (n=66) |
|---|---|---|
| **Were you absent from work?** | | |
| **Yes** | **59.5%** | **56.1%** |
| Unspecified | 47.0% | 45.5% |
| <7 days | 3.7% | 1.5% |
| 7-13 days | 2.5% | |
| 14-28 days | 3.0% | 1.5% |
| >28 days | 3.3% | 7.8% |
| **No** | **40.5%** | **43.9%** |
| **Did you suffer loss of earnings?** | **n=700** | **n=70** |
| **Yes** | **54.3%** | **41.4%** |
